# Comparing different neuroimaging modalities for quantification of the cholinergic system in Parkinson’s disease

**DOI:** 10.64898/2026.07.15.26357522

**Authors:** E. d’Angremont, T.M. Marschall, R.J. Renken, I.E.C. Sommer

## Abstract

**Introduction:** Parkinson’s disease (PD) is a multifactorial disorder, affecting multiple neurotransmitter systems, including the cholinergic system. Cholinergic denervation is heterogeneous across patients and difficult to predict based on clinical presentation. In this study, we assessed the sensitivity of structural MRI (sMRI) and functional MRI (fMRI) to cholinergic degeneration related to PD and to cognitive functioning in PD. We compared our results to results from previously reported [^18^F]Fluoroethoxybenzovesamicol ([^18^F]FEOBV) PET imaging, which is considered the gold standard for cholinergic imaging.

**Methods:** 34 PD patients and 10 healthy controls underwent structural T1-weighted MRI. A subset of 14 patients and 9 controls also underwent resting-state fMRI. We extracted the bilateral volumes of the nucleus basalis of Meynert (NBM) from the sMRI images. Functional connectivity (FC) from the NBM to the cortex (NBM-FC) was determined using fMRI data. Principal component analysis (PCA) was applied to reduce the dimensionality of the NBM-FC images. We assessed performances for NBM-FC in distinguishing patients from controls using stepwise logistic regression. Similarly, NBM volume was used using logistic regression. Furthermore, the relation between these measures and cognitive function in several domains was investigated with (stepwise) linear regression. Leave-one-out cross validation (LOOCV) and bootstrapping was performed to assess robustness of the results.

**Results:** NBM-FC was well able to discriminate patients from controls with an AUC of 0.84 (95% CI: 0.62-1). NBM volume showed lower performance, but was still better than chance: AUC: 0.75 (95% CI: 0.57-0.93). Significant correlations were found between 1) cognition in the attentional domain and NBM-FC (*r*=0.63; p=.015) and 2) global cognition and NBM volume (*r*=0.55, p=.001). These results were inferior to those previously reported using [^18^F]FEOBV tracer uptake (see Chapter 6). Bootstrapping revealed that NBM volume of only the left hemisphere was stably related to PD diagnosis and global cognition in PD patients. We found that a lower NBM-FC in specific brain areas, including the fusiform gyrus, supramarginal gyrus and dorsolateral prefrontal cortex, was related to PD diagnosis. Bootstrapping revealed no stable NBM-FC pattern related to attention.

**Conclusion:** Although MRI results were slightly inferior to [18F]FEOBV PET data, MRI may provide a cheaper and more widely available alternative for cholinergic imaging. We recommend testing the utility of MRI as predictor and monitor of cholinergic treatment effect in a longitudinal study.

## Introduction

Parkinson’s disease (PD) is the second most prevalent neurodegenerative disorder worldwide, with increasing age-standardized prevalence rates (1). Even though PD is often regarded as neurodegenerative disease that primarily affects the dopaminergic system, it is well recognized that PD is a multifactorial disorder, affecting multiple neurotransmitter systems, including serotonin, norepinephrine and acetylcholine. Studies even found a more severe cholinergic deficiency in PD compared to Alzheimer’s disease (AD), which is traditionally viewed as a primarily cholinergic disease (2). Acetylcholine is involved in numerous brain functions, including attention, memory and learning. Cholinergic degeneration, as observed in PD, has been related to cognitive deterioration, olfactory dysfunction, postural instability, gait disorders and visual hallucinations, among other symptoms (3). Cholinergic deficits can be partly compensated with cholinesterase inhibitors; however, these drugs have the potential to exacerbate motor symptoms (4), limiting their routine prescription in PD. Accurate assessment of cholinergic deficits in an individual patient would enable neurologists to tailor treatment strategies in accordance with personalized medicine principles.

Different neuroimaging modalities have been applied to investigate the cholinergic system in vivo, including PET and SPECT (3), as well as structural and functional MRI (5). [^18^F]Fluoroethoxybenzovesamicol ([^18^F]FEOBV) is a PET tracer that binds to vesicular acetylcholine transporter (VAChT). [^18^F]FEOBV PET is currently regarded the gold standard, as it corresponded well to gene expression and histological data in post-mortem brain tissue (6). Recent findings from our group show that [^18^F]FEOBV PET can reliably distinguish patients from controls and can also predict cognitive functioning within certain domains in PD patients (see Chapter 6).

However, [^18^F]FEOBV PET has some major drawbacks, such as the patient burden (radiation and intravenous tracer administration), its high cost and relatively low availability. A less invasive and more broadly available technique, such as MRI, would be helpful for clinical use. Both structural and functional MRI may provide valuable information about cholinergic system integrity. For example, volumetric analysis of the nucleus basalis of Meynert (NBM), the major source of cholinergic projections throughout the entire cerebral cortex and hippocampus, has shown differences between healthy controls and patients with PD related dementia (PDD) (7), and could predict future, but not current, cognitive deficits (8). In an earlier study, global cognitive functioning was correlated with volume of the substantia innominate (SI), a broader area in the basal forebrain including NBM among other brain structures (9). Studies applying resting-state functional MRI (fMRI) to calculate functional connectivity (FC) from the NBM to the cortex (NBM-FC) observed differences in posterior brain areas in the PD group (7), which were more pronounced in patients with cognitive impairment (10).

The sensitivity of volume of the NBM and NBM-FC to cholinergic degeneration in PD in comparison to [^18^F]FEOBV is currently unknown. The aim of this study is therefore to extract cholinergic measures from both structural and functional MRI and assess those in their ability to distinguish patients from controls and in their relation to cognitive functioning in PD. We compared the results to those from previously reported [18F]FEOBV PET imaging (Chapter 6).

## Methods

### Subjects

Thirty-four patients (twenty-six males) with a clinical diagnosis of PD and ten healthy controls (HC; eight males) underwent [^18^F]FEOBV PET imaging and a T1-weighted MRI scan in the University Medical Center Groningen. A subset of fourteen patients (ten males) and nine healthy controls (eight males) also underwent a resting-state functional MRI scan. Cognitive assessment was performed as described previously (see Chapter 6). In short, four different domains, i.e. attention, visuospatial function, executive function and memory, were assessed with at least two neuropsychological tests per domain (Table 1, Chapter 6). A z-score was calculated for each test, relative to the HC group. The z-scores were averaged within domains to form a composite z-score per domain. The average composite score was used as a measure for global cognition.

**Table 1.** Results of logistic regression analysis predicting disease state for each imaging modality.

|  | AUC (95% CI) |
| --- | --- |
| NBM volume | 0.75 (0.57-0.93) |
| NBM-FC | 0.84 (0.62-1) |
| [<sup>18</sup>F]FEOBV<br>SUVR | 0.91 (0.83-1) |

Performance of [<sup>18</sup>F]FEOBV SUVR was reported previously (Chapter 6) and is provided here for direct comparison. For NBM-FC, the results are based on a sub-sample of 14 patients and 9 controls (34 patients and 10 controls for the other measures). AUC = area under the receiver operating characteristic curve, NBM = nucleus basalis of Meynert, NBM-FC = functional connectivity from the NBM, SUVR = standardized uptake value ratio.

### Imaging acquisition and processing

#### Structural MRI

Structural T1-weighted images were acquired with a Siemens Prisma 3T scanner, with a magnetization-prepared rapid gradient-echo (MPRAGE) sequence, 2.98 ms echo time, 2300 ms repetition time, 900 ms inversion time, 9° flip angle, 256 mm field of view, yielding a voxel size of 1.0 × 1.0 × 1.2 mm^3^.

The T1-weighted images were segmented into gray and white matter images and spatially normalized utilizing the CAT12 toolbox (11) of Statistical Parametric Mapping (SPM version 12, Wellcome Trust Center for Neuroimaging), with default settings. The spatial normalization was visually checked. The resulting gray matter images were smoothed with an 8×8×8 mm kernel, after which the mean values of the bilateral NBM were extracted, using the Julich brain atlas (12) for delineation of the NBM. The CAT12 toolbox was also used to extract the total intracranial volume (TIV) of each subject. The NBM volumes were subsequently corrected for TIV by a Gram-Schmidt orthogonalization (13).

#### Functional MRI

Resting-state scans of 10 minutes (n = 300 volumes) were obtained with the same Siemens Prisma 3T scanner, using a T2*-weighted multi-echo EPI (echo planar imaging) sequence with echo times 9.74 ms, 20.91 ms and 32.08 ms, 37 axial slices, anterior–posterior phase encoding, voxel size of 3.5 mm^3^, 2000 ms repetition time, and a field of view of 256×249 mm. Participants were instructed to keep their eyes closed, let their mind roam freely and not fall asleep.

Preprocessing of the fMRI data was conducted using fMRIPrep 22.0.2 (14). Anatomical T1-weighted images were corrected for intensity non-uniformity and skull stripped. These scans were normalized to MNI space and used for anatomical reference. The following steps were applied to all functional BOLD images: (1) generation of a skull stripped reference volume based on the shortest echo, (2) head-motion correction, (3) slice -timing correction. The separate echoes were combined and denoised with tedana (15). Following, the combined images were coregistered and normalized to MNI space using ANTs (http://picsl.upenn.edu/software/ants/).

Hereafter, a seed to voxel connectivity analysis was performed in Conn (16). For this, data were filtered with a band pass filter (0.008-0.09Hz). Fisher-transformed bivariate correlation coefficients, i.e. functional connectivity (FC), were calculated between all voxels and the bilateral NBM, again applying the Julich brain atlas for delineation of the NBM. The resulting images were smoothed with an 8×8×8 mm kernel. As the NBM projects to the cerebral cortex, we applied a cortical mask, including hippocampus and amygdala, which was created with the neuromorphometrics atlas (http://www.neuromorphometrics.com/) and also smoothed with an 8×8×8 mm kernel. To further reduce dimensionality of the final images, we performed principal component analysis (PCA) with >90% of the total variance retained. The resulting principal components (PCs) were used for further analysis.

#### [^18^F]FEOBV PET

Brain [^18^F]FEOBV PET scans were obtained and processed as described previously (Chapter 6). In short, subjects underwent a low-dose CT scan followed by a 30 minute PET scan, starting 210 minutes after injection of approximately 200 MBq [^18^F]FEOBV. The images were co-registered with individual T1-weighted MRI scans and subsequently intensity normalized, yielding parametric standardized uptake value ratio (SUVr) images. For reference region, we employed an eroded version of the supratentorial white matter mask, derived from a FreeSurfer (http://surfer.nmr.mgh.harvard.edu/) segmentation of the T1-weighted MRI scan, as previously described (17). Subsequently, parametric SUVr images were corrected for partial volume effects (PVE) using the Muller-Gartner method (18). The images were subjected to spatial normalization into the MNI space, followed by smoothing using an 8×8×8 mm kernel. Per subject, a mask was created based on relative thresholding, including ≥3% of the maximum SUVr value. Hereafter, we created a global mask including only voxels that were present in the masks of each individual subject. The global mask was applied to all normalized SUVr images. Dimensionality of the PET images was reduced with PCA, retaining >90% of total variance.

### Prediction of disease state

For NBM-FC, the same process was followed as for SUVr om Chapter 6: the PCs were utilized as independent variables in a stepwise logistic regression model to predict disease state (‘PD’ or ‘HC’). The Bayesian information criterion (BIC) was used for determination of the optimal selection of PCs. For the volumetric NBM analysis, the TIV-corrected volumes of the left and the right NBM were used as independent variables in a regular logistic regression model. We applied a leave-one-out cross validation (LOOCV), where in each iteration a (stepwise) logistic regression model was fitted on all data minus the participant left out and subsequently applied on the left-out participant. Hereafter, we calculated the area under the curve (AUC) of the receiver operating characteristic (ROC) to compare performances between measures. To calculate the 95% CI of each AUC and statistically compare performance between different modalities, we applied the method described by DeLong et al. (19), as implemented in the pROC package in R (20).

### Prediction of cognitive functioning

Similarly, we fitted linear regression models to predict cognitive functioning in different domains within the PD group with LOOCV. For NBM-FC and SUVr, stepwise linear regression models were fitted in each iteration with the PCs as independent variables and the cognition z-score as dependent variable, again using BIC. For the volumetric analysis, the TIV-corrected volumes of the left and right NBM were chosen as independent variables in a regular linear regression model. We calculated the Pearson’s correlation between the predicted z-scores based on the left-out participant and the true z-scores to assess performance of the different modalities.

### Bootstrapping

To assess the stability of the different modalities in prediction of disease state and cognitive functioning, we performed bootstrapping analysis with 5000 repetitions. In each repetition, again a (stepwise) logistic or linear regression model was fitted on a random sample. In disease prediction, the number of PD and HC subjects was kept constant in each iteration. For NBM volume, we considered the relation to PD reliable if the 95% CI of the model coefficients did not straddle zero. For NBM-FC and SUVr, in each repetition a voxel weight was determined by a weighted sum of the stepwise logistic regression model coefficients and the eigenvectors of the corresponding PCs. To visualize the resulting brain patterns, we selected the median value for each voxel and excluded voxels for which the 95% CI straddled zero.

## Results

Demographics and cognition scores of the included subjects are shown in Table 2 of Chapter 6. There were no significant differences between groups in age, sex, or educational level. The PD group had significantly lower cognition scores for all domains, with the exception of memory. Demographics and cognition scores for the fMRI subsample are shown in Supplementary Table 1. In this subsample, a significant difference was found only for visuospatial cognition.

**Table 2.** Correlations between cognitive functioning in different domains and predicted values based on linear regression analysis.

| Cognition pattern | [$^{18}\text{F}$]FEOBV SUVr | | NBM volume | | NBM-FC | |
| --- | --- | --- | --- | --- | --- | --- |
|  | *r* | p-value | *r* | p-value | *r* | p-value |
| Attention | 0.36 | .036 | -0.24 | .173 | 0.63 | .015 |
| Executive function | 0.39 | .022 | 0.25 | .161 | - | - |
| Memory | 0.16 | .364 | -0.01 | .934 | - | - |
| Visuospatial cognition | 0.55 | .001 | 0.11 | .543 | - | - |
| Global cognition | 0.50 | .003 | 0.55 | .001 | 0.09 | .748 |

Correlations for SUVr were reported previously (Chapter 6) and are provided here for direct comparison. For NBM-FC, the results are based on a sub-sample of 14 patients and 9 controls (34 patients and 10 controls for the other measures). NBM = nucleus basalis of Meynert, , NBM-FC = functional connectivity from NBM, SUVr = standardized uptake value ratio.

### Prediction of disease state

NBM volume and NBM-FC respectively showed a fair and considerable performance in distinguishing patients from controls (Table 1) (see (21) for the interpretation of AUC values). For both measures the AUC 95% CI straddled 0.50, indicating that they performed significantly better than chance. The AUC values of both MRI measures were inferior to that of global [^18^F]FEOBV SUVr; however, there were no significant differences between pairs of AUCs. The ROC curves of all modalities are shown in Figure 1. The performance and ROC curves of all modalities based on the subsample of participants with fMRI are shown in Supplementary Table 2 and Supplementary Figure 1, respectively.

**Figure 1.**
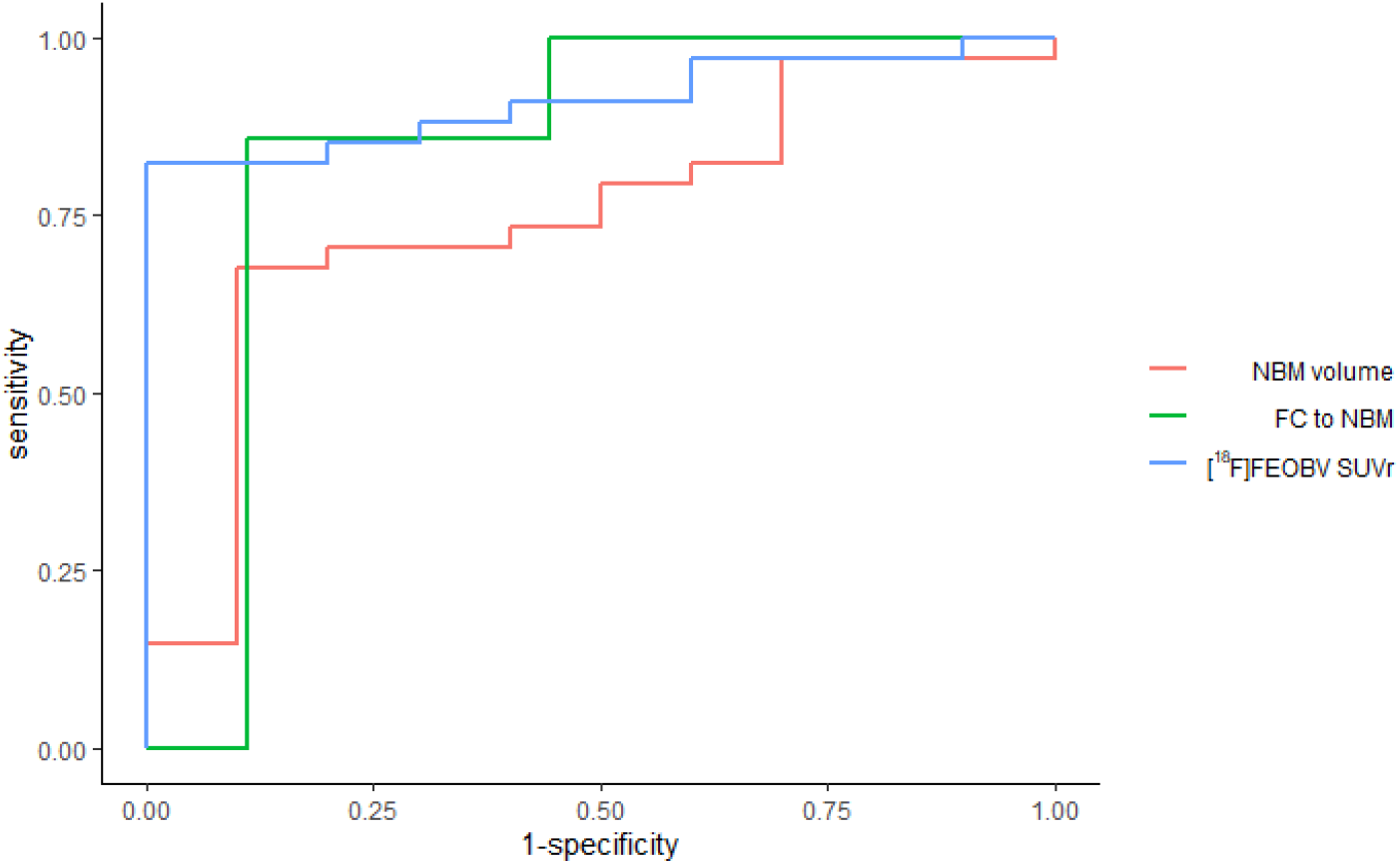
Receiver operating characteristic (ROC) curves per imaging modality resulting from a leave-one-out cross validation of disease prediction using logistic regression. For NBM volume and [^18^F]FEOBV SUVr, the results are based on a sample of 34 patients and 10 controls. For NBM-FC, the results are based on a sub-sample of 14 patients and 9 controls (34 patients and 10 controls for the other measures). The ROC curve for SUVr was reported previously (Chapter 6) and is provided here for direct comparison. NBM = nucleus basalis of Meynert, FC = functional connectivity, SUVr = standardized uptake value ratio.

Bootstrapping analysis showed that NBM volume in the left hemisphere was reliably related to PD diagnosis (median coefficient: -75.2; 95% CI: -394.5 to -8.2). The negative value indicates that a smaller volume was associated with PD diagnosis. NBM volume in the right hemisphere, however, showed an unstable relation to PD (median coefficient: 56.0; 95% CI: - 49.4 to 343.8). Figure 2 shows the brain pattern of the relation between NBM-FC and PD, after bootstrapping, alongside the pattern of [^18^F]FEOBV tracer uptake. Compared to the [^18^F]FEOBV PET pattern, the pattern of NBM-FC appeared to be more confined to specific brain areas, particularly the bilateral fusiform gyrus, bilateral supramarginal gyrus, left precentral gyrus and bilateral dorsolateral prefrontal cortex (DLPFC). Relatively small clusters with a stable positive relation to PD were found, mainly in the frontal lobe, right temporal lobe and one in the anterior cingulate gyrus. The SUVr pattern based on the subsample of participants with fMRI is shown in Supplementary Figure 2.

**Figure 2.**
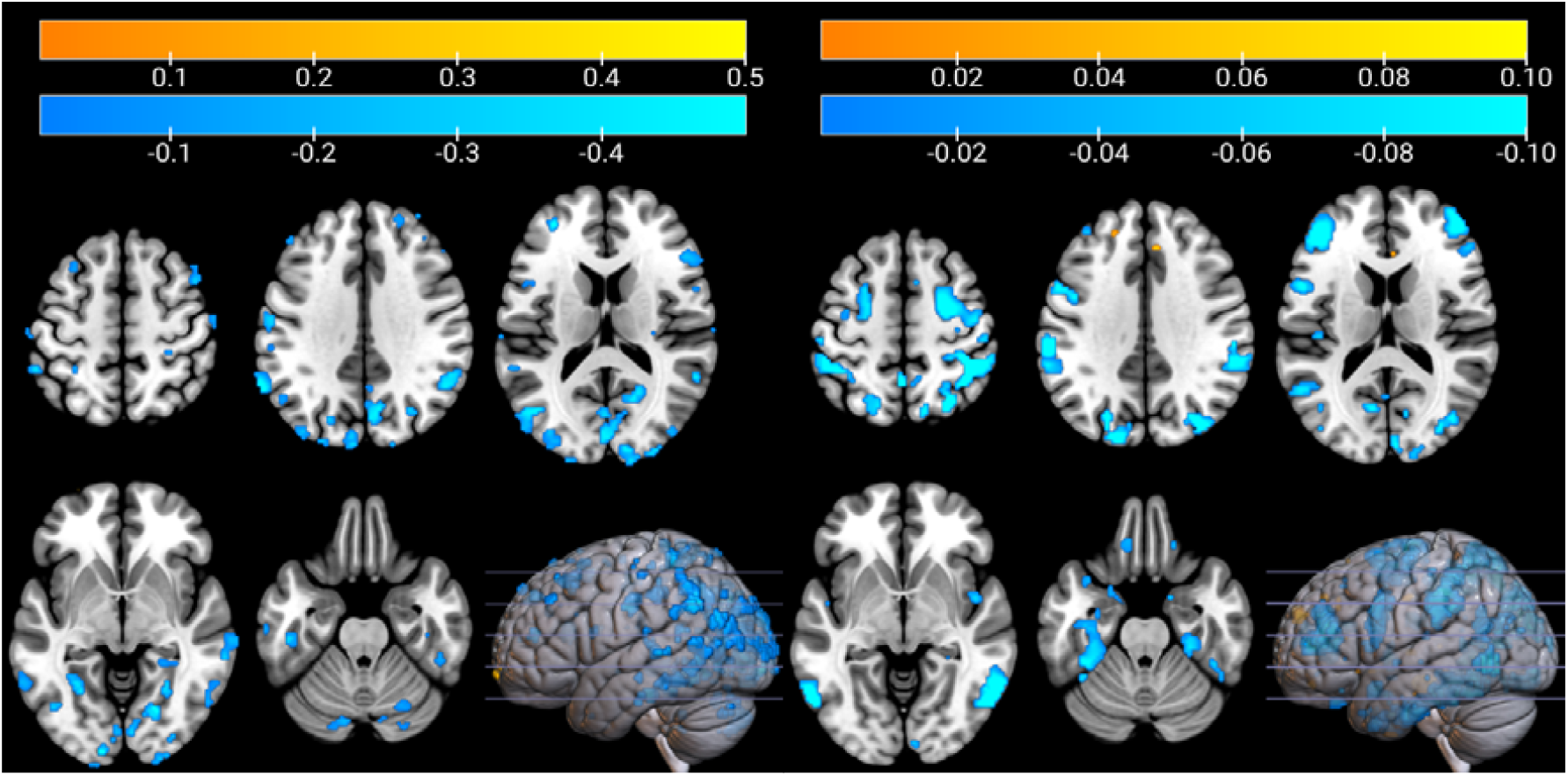
Stable pattern weights (i.e. coefficients) of [^18^F]FEOBV tracer uptake (left) and functional connectivity to the nucleus basalis of Meynert (right) after bootstrapping. A negative value means that a decrease in tracer uptake or connectivity is related to Parkinson’s disease. The pattern for tracer uptake was reported previously (Chapter 6) and is provided here for direct comparison. For NBM-FC, the results are based on a sub-sample of 14 patients and 9 controls (34 patients and 10 controls for [^18^F]FEOBV PET).

### Prediction of cognitive functioning

Table 2 shows the correlations between the cognition z-scores of different domains and the predicted z-scores of the left-out participant in the LOOCV. Predicted values based on NBM volume showed a significant correlation with global cognition (Figure 3A). NBM-FC could predict attention with a significant correlation (Figure 3B), but for executive functioning, memory and visuospatial cognition, the stepwise regression model did not select any of the PCs. This means that the predicted scores were solely based on the intercept, so no correlations are presented for these domains. As reported previously, [^18^F]FEOBV PET was able to predict cognitive functioning of all included cognitive domains except memory with a significant correlation.

**Figure 3.**
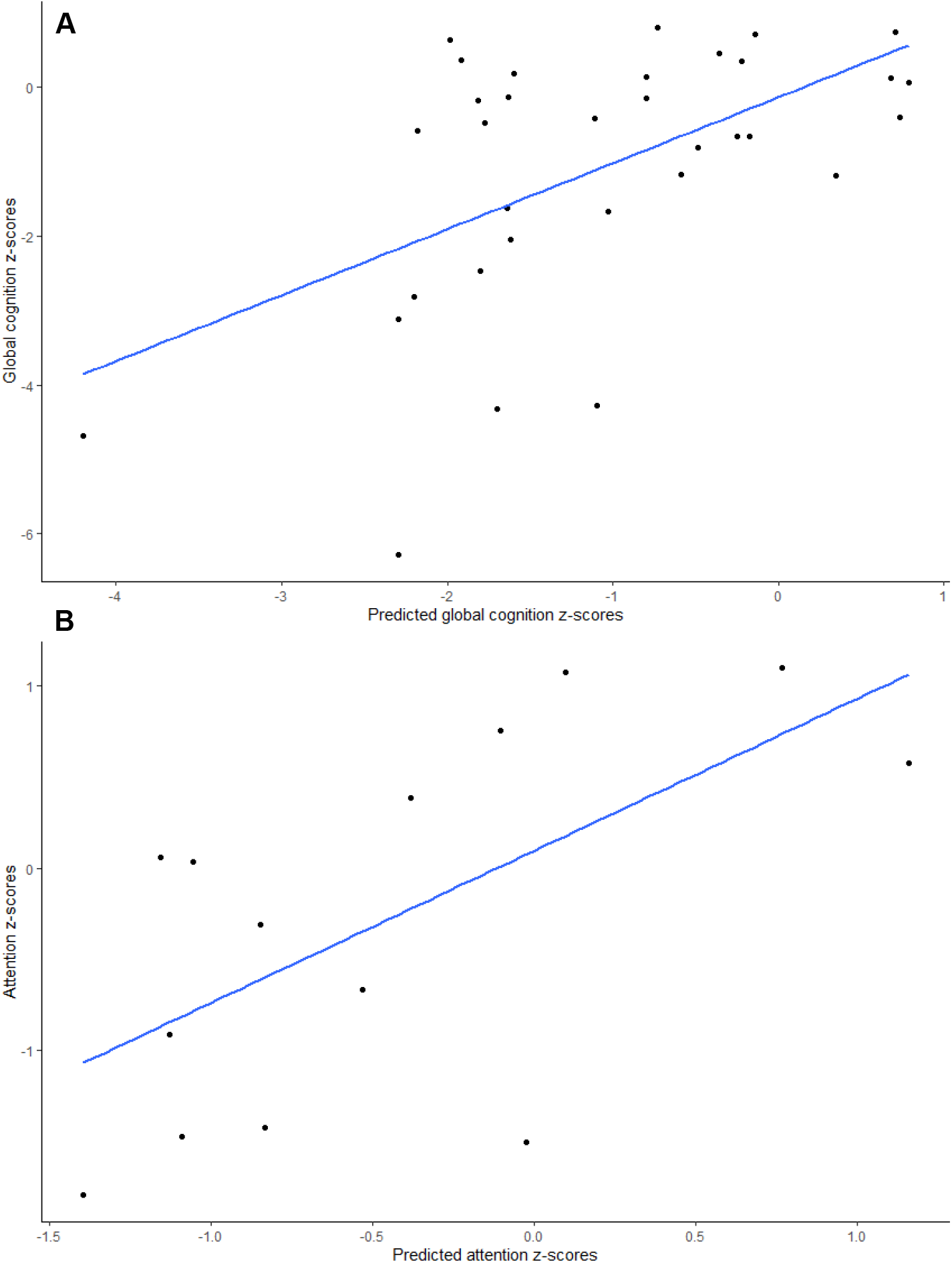
A: correlation between global cognition z-scores and predicted scores based on volume of nucleus basalis of Meynert (NBM). B: correlation between attention z-scores and predicted scores based on functional connectivity from the NBM. The predicted scores of both figures are the result of leave-one-out cross validation.

Although NBM-FC produced a significant correlation in the LOOCV with attention, this did not result in a stable pattern in the bootstrapping analysis. The other cognitive domains, as well as global cognition, neither produced stable patterns. Only left NBM volume was reliably related to global cognition after bootstrapping (median coefficient: 28.4; 95% CI: 3.4 to 54.3), with a smaller volume related to lower cognitive functioning. In all individual cognitive domains, there was no stable relationship with NBM volume.

## Discussion

This is the first study to compare three different measures for cholinergic imaging in PD. We found that both structural and functional MRI, in the form of NBM volume and NBM-FC, respectively, were able to distinguish between patents and controls with above chance performance. NBM volume could predict global cognition in PD and NBM-FC was specifically related to attention. However, both measures were inferior to previously reported results using [^18^F]FEOBV PET imaging, which showed excellent discriminatory ability and was also able to predict cognitive functioning across multiple domains in PD. Nonetheless, MRI may provide a cheaper and more widely available alternative to cholinergic PET imaging.

### Prediction of disease state

NBM-FC performed reasonably well in distinguishing patients from controls with an AUC of 0.84, which can be interpreted as ‘considerable’ (21). It should be noted, however, that fMRI was only performed in a subsample of participants, and [^18^F]FEOBV SUVr was able to achieve a perfect prediction in this subsample (Supplementary Table 2), suggesting that the subsample included some of the more advanced stage patients. NBM volume could also distinguish patients from controls significantly better than chance, with an AUC of 0.75, which can be interpreted as ‘fair’ (21). This is in line with previous findings where there was a significant difference in NBM volume between patients with Lewy body dementia and healthy controls, but with considerable overlap (7). Interestingly, specifically NBM volume of the left hemisphere showed a robust relationship to PD diagnosis in our results. It is unknown whether this finding has any clinical significance.

Where the SUVr-based PD-related pattern was relatively scattered across posterior brain regions and some superior frontal areas, the pattern of impaired NBM-FC related to PD diagnosis appeared to follow anatomically defined brain regions. The differences in patterns may reflect the different aspects of the cholinergic system that are assessed with both modalities. The FC pattern included regions associated with visual processing and also regions within the frontoparietal network, specifically the DLPFC and inferior parietal cortex. Dysfunction of the frontoparietal network has been related to impaired top-down control of attention in PDD (22).

### Prediction of cognitive functioning

Attention was the only cognitive domain that NBM-FC was related to in the LOOCV. Although the correlation was relatively strong, bootstrapping did not result in a stable pattern for this domain, suggesting that there is a strong between-subject variability in the cholinergic-specific functional network for attention. The failure of NBM-FC to predict cognitive functioning in other domains may be attributed to the relatively small sample size, being a subset of our total number of included participants. Alternatively, it is possible that NBM-FC is specifically related to attention, which fits with the idea of attention being the main function of cholinergic neuromodulation, through which other symptoms, such as deficits in other cognitive domains or visual hallucinations, may occur (23,24). VAChT expression, on the other hand, appeared to be more generally related to cognition, with largely overlapping patterns for attentional, executive and visuospatial functioning. Alternative approaches for fMRI analysis may be assessed in the future, such as quantification of FC within brain networks known to be related to cholinergic activity, including the default mode network and the salience network (25), instead of using NBM as seed region.

NBM volume was related to global cognition, which is in line with previous research, showing a correlation between SI volume and Mini Mental State Examination score (9). However, NBM volume appeared to be less sensitive to functioning within specific cognitive domains. A possible explanation may lie in the fact that NBM volume, unlike the other two measures, merely evaluates the somas of the cholinergic nerves, located in the NBM, while there is mounting evidence that Lewy body deposition in PD pathology predominantly starts in axons, rather than cell bodies (see (26), for an overview). This phenomenon, termed ‘dying back’ degeneration, would make it probable that fMRI and PET are more sensitive to cholinergic denervation in PD, also from a theoretical point of view. Volumetric analysis of the NBM would only be able to identify a volume loss after a certain number of cell bodies have died. Moreover, in a mouse model of AD volume loss in the basal forebrain did not correlate with the reduction in number of cholinergic neurons, making its use as cholinergic biomarker debatable (27). More advanced MRI measures of the NBM, such as free water fraction, may be more sensitive to cognitive deterioration (28).

[^18^F]FEOBV is the most established method for cholinergic imaging in current literature, with thorough validation including correlation with regional gene expression levels and postmortem histological data (6). It is therefore not surprising that this method appears as the most reliable method in our results. However, as mentioned in the introduction, PET imaging has several downsides, such as its costs, the use of radiation and limited availability, whereas MRI is a much faster procedure, with lower costs and lower patient burden. Therefore, it may be worthwhile to investigate whether MRI analysis could be further optimized to increase sensitivity to cholinergic changes. Moreover, the reported performance of MRI as a cholinergic biomarker may already be sufficient for clinical purposes. Clinical utility for prediction and monitoring of cholinergic treatment effect should be assessed in a longitudinal study.

Additionally, it should be noted that the different measures reported here assess different aspects of the cholinergic system, which may be complementary in providing a comprehensive picture of cholinergic degeneration in an individual patient.

### Limitations

For several patients, extensive fMRI scans were prohibited by the presence of metal objects in the body that could not be removed. Therefore, the fMRI results in this study were based on a smaller subsample of participants, which limits the reliability of these findings and makes it more difficult to compare to the other two measures.

Additionally, the sample size of our control group is relatively small, making the constructed z-scores difficult to interpret.

## Conclusion

Performance of functional and structural MRI as a cholinergic biomarker in PD was inferior to that of [^18^F]FEOBV PET, both in the prediction of disease state as in the prediction of cognitive functioning. However, given the acceptable performance of structural and especially functional MRI and their practical advantages over PET, additional research should aim to assess the viability of MRI as a cholinergic biomarker in a longitudinal study, for example in prediction and monitoring of cholinergic treatment effect.

## Data Availability

All data produced in the present study are available upon reasonable request to the authors

## Supplementary materials

**Supplementary Table 1.**
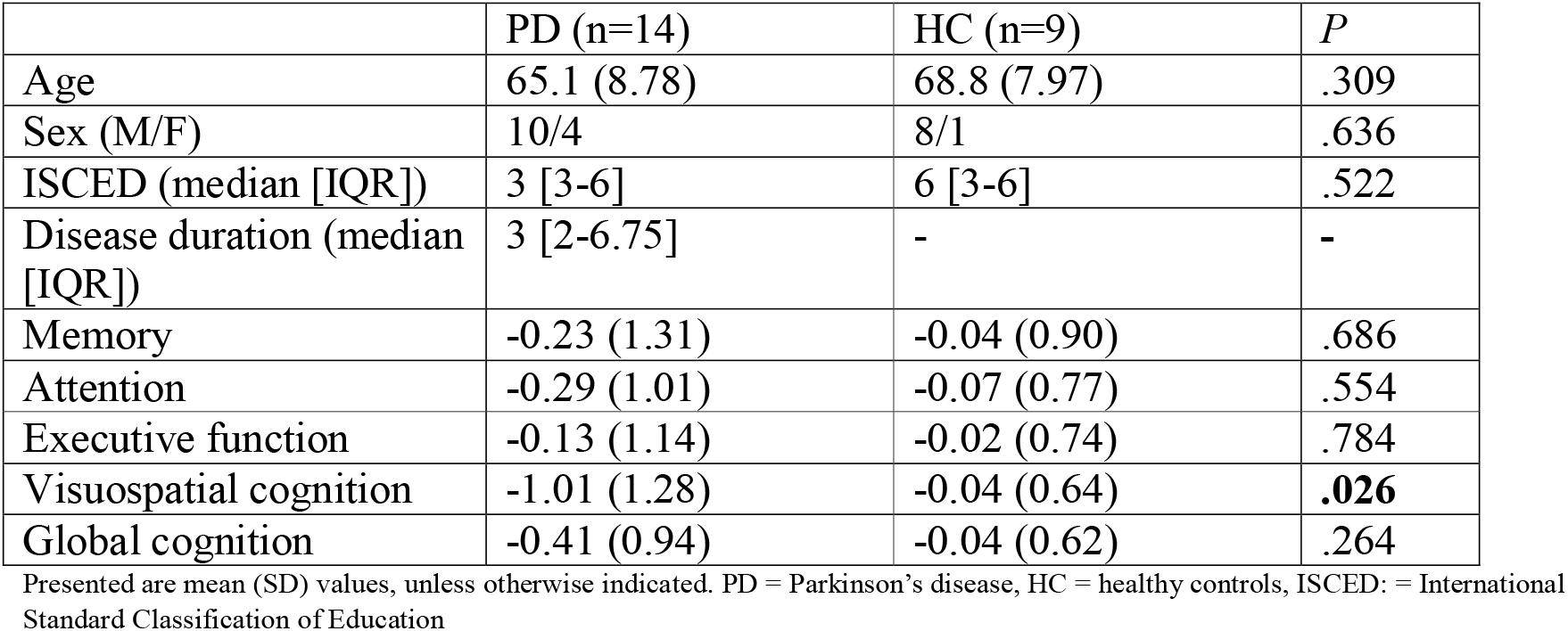
Demographics and cognition scores of subsample of participants with functional MRI.

**Supplementary Table 2.** Results of logistic regression analysis predicting disease state for each imaging modality in the subsample of participants with functional MRI.

|  | AUC (95% CI) |
| --- | --- |
| NBM volume | 0.64 (0.39-0.90) |
| FC to NBM | 0.84 (0.62-1) |
| [<sup>18</sup>F]FEOBV SUVr | 1 (1-1)* |

AUC = area under the receiver operating characteristic curve, NBM = nucleus basalis of Meynert, FC = functional connectivity, SUVr = standardized uptake value ratio. \*Significant difference with NBM volume (Z=-2.77, P=.006), but 95% CI may be misleading with an AUC of 1.

**Supplementary Figure 1.**
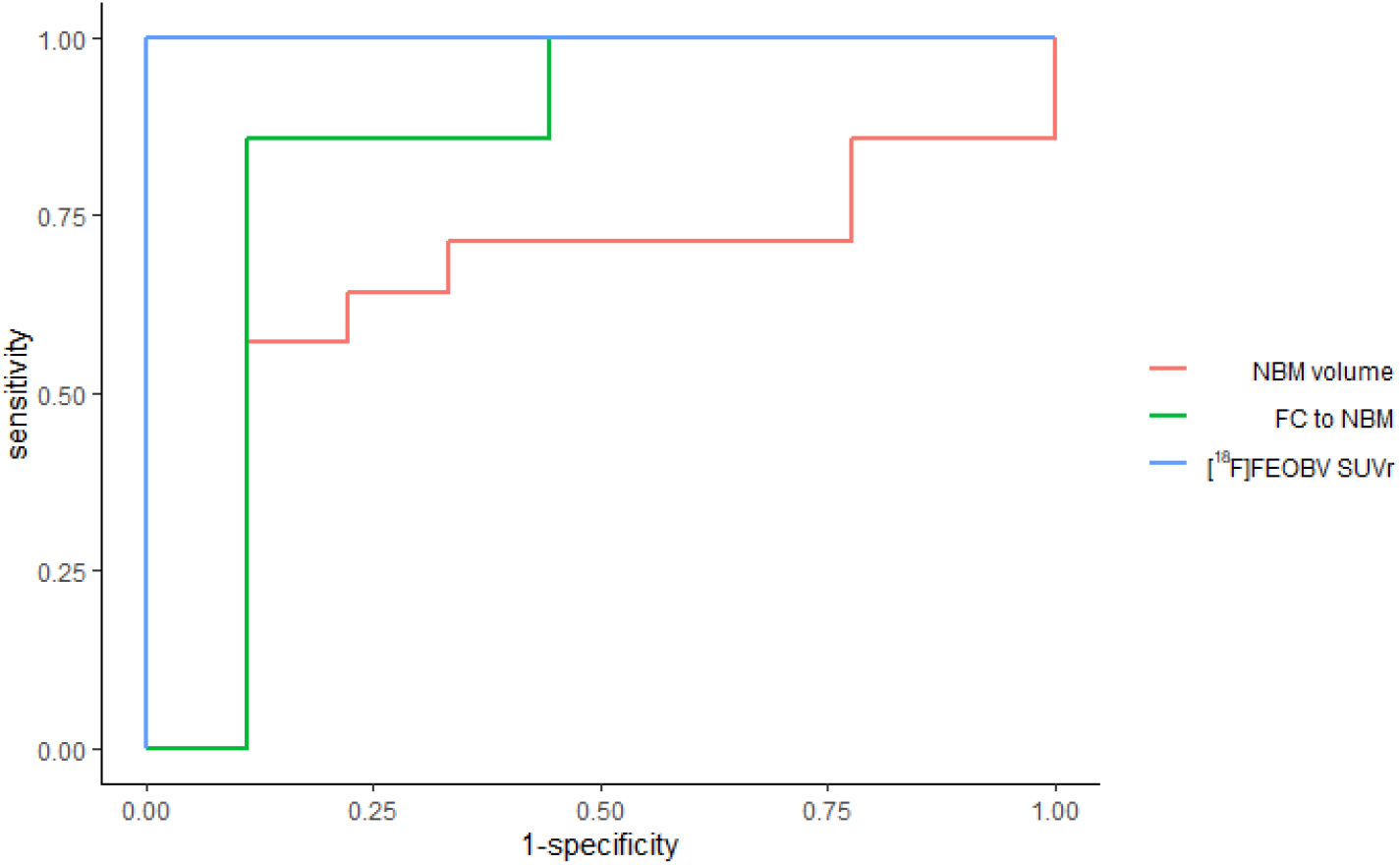
Receiver operating characteristic curves per imaging modality resulting from a leave-one-out cross validation of disease prediction using logistic regression. Results are based on a subsample of 14 patients and 9 controls. NBM = nucleus basalis of Meynert, FC = functional connectivity, SUVr = standardized uptake value ratio.

**Supplementary Figure 2.**
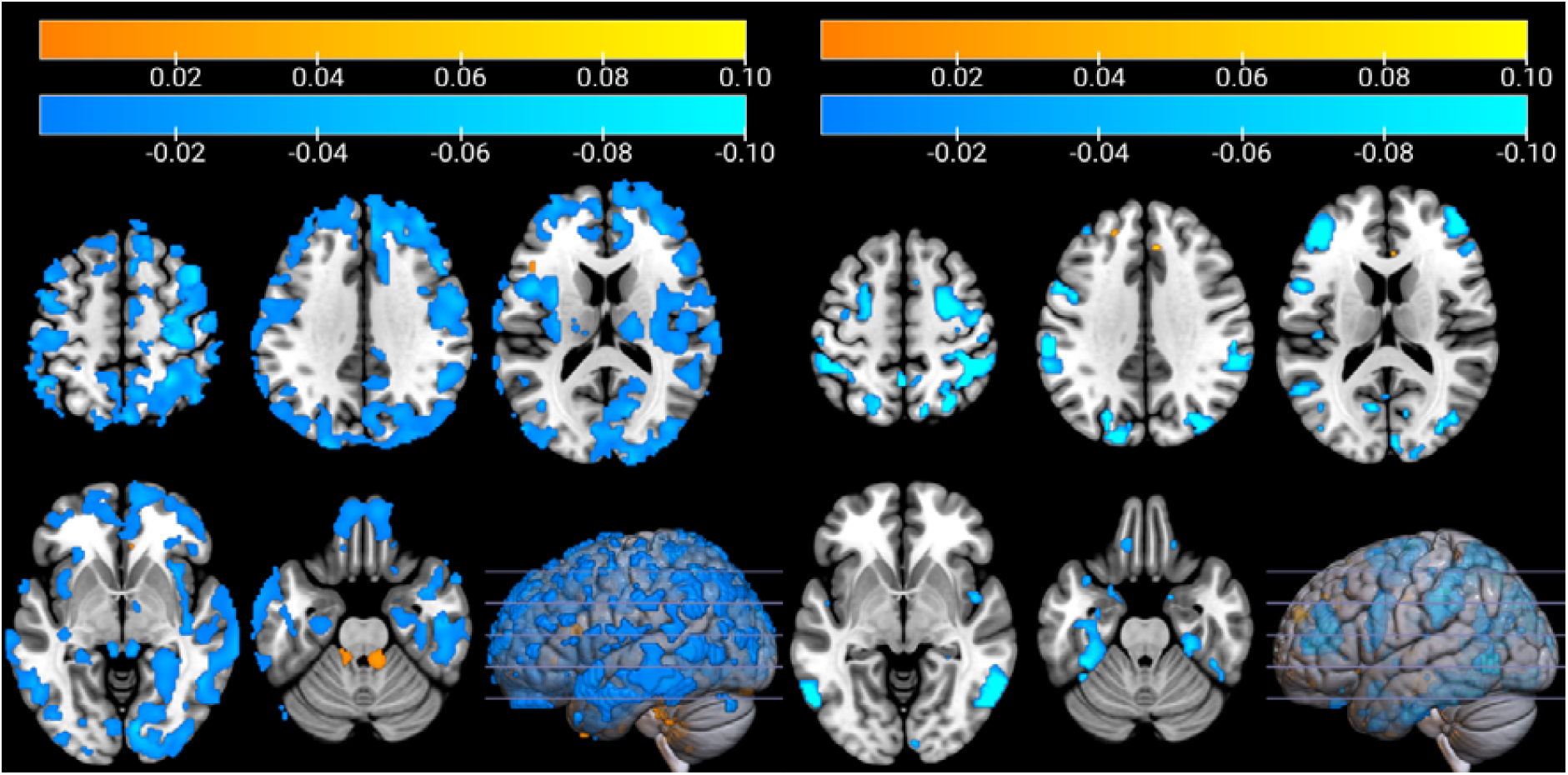
Stable pattern weights (i.e. coefficients) of [^18^F]FEOBV tracer uptake (left) and functional connectivity to the nucleus basalis of Meynert (right) after bootstrapping. A negative value means that a decrease in tracer uptake or connectivity is related to Parkinson’s disease. Results based on subsample of participants with functional MRI.

## Notes

### Competing Interest Statement

The authors have declared no competing interest.

### Author Declarations

The Ethics committee of University Medical Center Groningen gave ethical approval for this work.

### Funding

- ZonMw, The Dutch Organisation for knowledge and innovation in health, healthcare and well-being: 636310010

